# Toward systems-level analysis of medical incident reports: A clinician-annotated evaluation of large language models using HFACS

**DOI:** 10.64898/2026.11.24.26368854

**Authors:** Fuyuto Miyake, Takashi Watari, Shintaro Kosaka, Yudai Goto, Natsumi Edagawa, Tomohiro Takayama, Kentaro Ishida, Ryuhei So, Yuki Kataoka

## Abstract

**Objectives:** We aimed to assess agreement between reviewers in factor extraction and Human Factors Analysis and Classification System (HFACS) classification, and between large language model (LLM) classifications and reviewer-consensus reference standards, using public medical incident reports from Japan.

**Methods:** Six clinicians in three pairs reviewed 150 records with artificial intelligence (AI)-generated fault tree analysis (FTA) candidates and 60 new records without AI candidates, creating pair-consensus reference standards (RSv1 and RSv2). We assessed classification agreement, factor-extraction micro-F1, and FTA versus direct classification. Reviewers reassessed RSv2 after additional public information. AI classification was also evaluated on fixed RSv2 factors.

**Results:** Pooled inter-reviewer detailed-category kappa in RSv1 was 0.503 (95% CI, 0.456 to 0.548). In the final RSv1 evaluation (37 records; 306 leaves), kappa was 0.427 (0.359 to 0.493) for GPT-5.6 Sol and 0.499 (0.445 to 0.551) for secondary Kimi K2.6. Neither estimate reached the detailed-category kappa target of 0.6. FTA-based structural rankings and factor-frequency rankings selected identical top-five categories; the FTA-minus-direct-classification micro-F1 difference was 0.032 (−0.024 to 0.086). RSv2 factor-extraction micro-F1 was 0.420 (0.303 to 0.527), using AI-assisted semantic matching; 23/60 RSv2 records were analysable with uncertainty and 22/60 were not analysable. Responses without uncertainty accounted for 37.5% before and 46.7% after reassessment; pooled linear weighted kappa for inter-reviewer analysability agreement was 0.386 and 0.203. Exploratory AI classification of 75 RSv2 factors with a single HFACS label yielded detailed-category kappa 0.240 (0.101 to 0.390).

**Conclusions:** Disagreement persisted in the extraction and classification of contributing factors from public medical incident reports, including in reviewer assessments, and the tested FTA approach did not establish added value. Difficulty obtaining sufficiently specific contextual information from the available narratives may have contributed to these judgement difficulties, consistent with concerns raised in previous studies. Organisations that operate incident reporting systems or develop reporting guidance could strengthen reporting processes so that the specific circumstances and context needed to interpret contributing factors and relationships are recorded.

## 1. Introduction

Learning from medical incident reports to prevent recurrence involves analysing contributing factors and relationships, identifying targets for improvement, and implementing and evaluating preventive measures. These factors encompass not only individual actions at the point of care but also team working conditions, supervision and organisational context. An analysis of 126 serious incident investigation reports from a single NHS trust in England found an emphasis on individual actions with insufficient attention to underlying system and organisational factors [1]. Linking incident analyses to systems-level improvement therefore requires a common framework for organising factors from individual actions to supervisory and organisational conditions.

The Human Factors Analysis and Classification System (HFACS) provides one such framework. Based on the Swiss cheese model, it organises contributing factors into four levels: unsafe acts, preconditions for unsafe acts, unsafe supervision and organisational influences [2,3]. HFACS and its adaptations have been used to analyse contributing factors in surgical near-miss reports and serious incident investigation reports [1,4]. To support the analysis of large numbers of reports, large language model (LLM) classifications of contributing factors have also been evaluated against expert judgement [5,6]. Ebara et al. evaluated BERT-based sentence-level multilabel classification using a modified P-mSHELL framework on selected background descriptions from nationwide public medication-related incident reports [7]. However, classification of selected sentences or expert-extracted factor statements does not establish whether reviewers can identify the same specific contributing factors from reports. To use report analysis to identify improvement targets at individual, team and organisational levels, it remains insufficiently understood how consistently reviewers extract factors and classify them using HFACS in public reports not restricted to a single clinical specialty, and how closely LLM classifications agree with reviewer-consensus reference standards derived from these assessments.

We therefore aimed to assess agreement between reviewers in factor extraction and HFACS classification, and between LLM classifications and reviewer-consensus reference standards, using public medical incident reports collected from healthcare institutions across Japan without restriction to a single clinical specialty.

## 2. Methods

### 2.1. Study design and data source

We conducted a cross-sectional study of public medical incident records. We assessed agreement along two axes: (i) agreement between independent human reviewers, and (ii) agreement between artificial intelligence (AI) classifications and reference standards created by reviewer-pair consensus (RSv1 and RSv2). We report this study following the Guidelines for Reporting Reliability and Agreement Studies (GRRAS) checklist [8] (Supplementary Checklist).

Figure 1 summarises the staged evaluation using two separate cohorts of public medical incident records. In the first cohort (150 records), reviewers independently classified AI-generated candidate factors and created a reference standard through pair consensus (RSv1). We used this cohort to assess classification agreement and the added value of fault tree analysis (FTA). In a separate cohort (60 new records), reviewers independently extracted and classified factors without AI-generated candidates and created a second reference standard through pair consensus (RSv2). We assessed extraction and classification agreement, reassessed analysability and HFACS classification with additional public information, and performed exploratory AI classification of the RSv2 factors.

**Figure 1.**
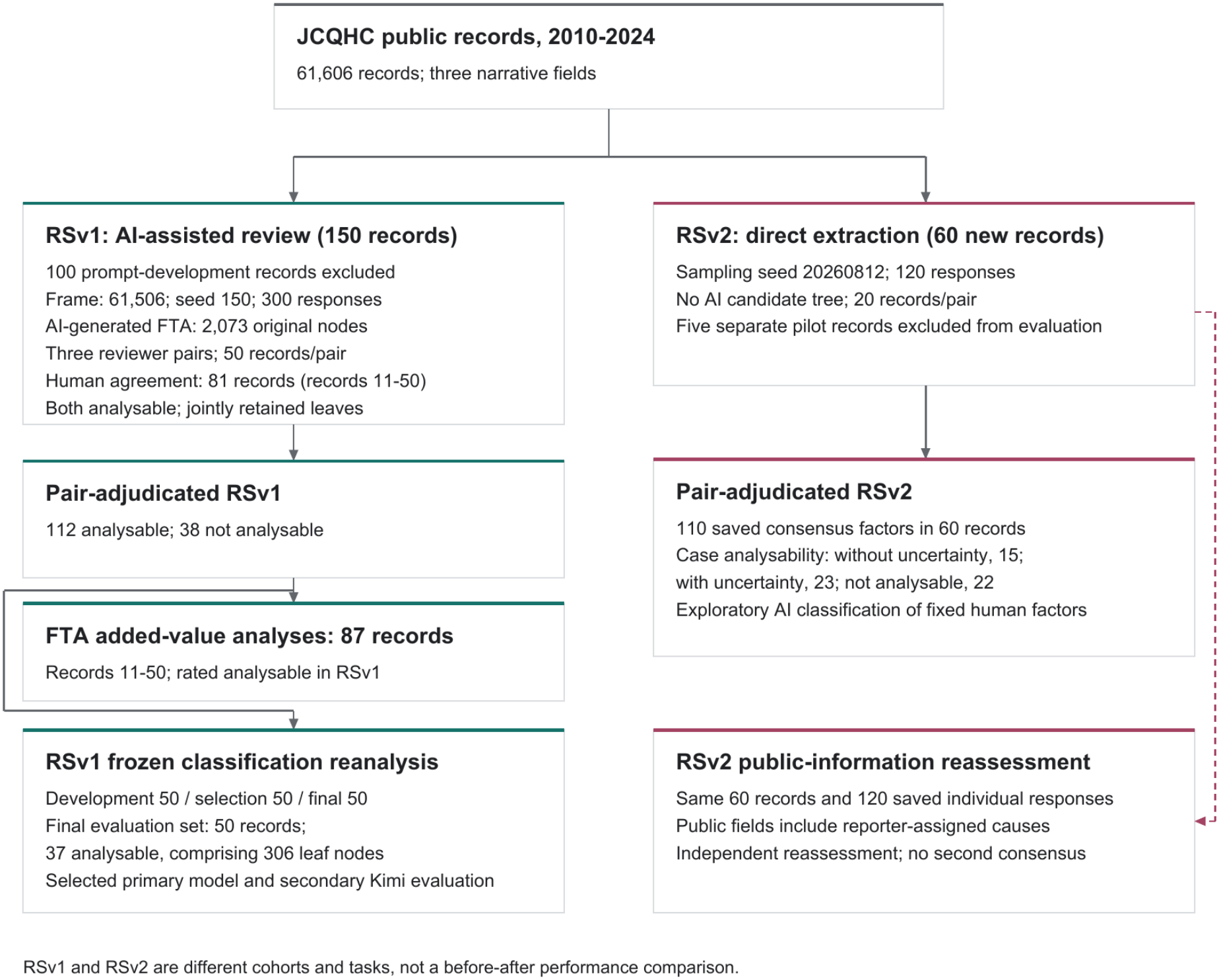
Study flow and distinct evaluation cohorts. The RSv1 cohort included 150 public records reviewed using AI-generated fault tree analysis (FTA) candidates. Among records 11–50 assigned to each pair, inter-reviewer classification included 81 records considered analysable by both reviewers; FTA added-value analyses included 87 records rated analysable in RSv1. The RSv2 cohort involved independent human factor extraction without AI-generated candidates in 60 new records, following a separate five-record pilot. The dashed path carries independent responses into public-information reassessment, not RSv2. The RSv1 and RSv2 cohorts involved different records and tasks; differences between their scores should not be interpreted as performance improvement. The 50/50/50 classification split was a later reanalysis, not an untouched external cohort. AI, artificial intelligence; JCQHC, Japan Council for Quality Health Care; RS, reference standard.

The source database was the public case database of the Project to Collect Medical Near-Miss/Adverse Event Information, operated by the Japan Council for Quality Health Care (JCQHC). The project collects adverse event reports through a web-based reporting system from institutions subject to mandatory reporting and voluntary participants, including hospitals and clinics [9,10]. As of 31 March 2025, 4,176 institutions participated in its adverse event reporting component: 270 mandatory-reporting institutions, mainly university hospitals and national hospitals, and 3,906 voluntary hospitals and clinics [11]. Public records contain information submitted by medical institutions at the time of reporting; JCQHC removes information that could identify individuals or institutions before publication [12]. The sampling frame comprised 61,606 unique public medical incident records from 2010–2024, excluding near-miss records. We analysed the event description, background, and improvement-measure fields of these public records, rather than internal hospital incident reports or medical records. The evaluation was therefore limited to information reported in these fields; the underlying events were not independently verified against hospital records. Additional public fields were used in the reassessment described in Section 2.6.

For RSv1, we randomly sampled 150 records from the sampling frame after excluding the 100 records used for prompt development and allocated 50 records to each of three reviewer pairs. For RSv2, we selected 60 new records using a fixed, reproducible procedure after excluding previously used records and pilot candidates, allocating 20 records to each pair. We determined the number of records for manual review with reference to previous agreement studies of patient safety incident analysis, which used 50 to 150 reports [13,14]. Accordingly, we set the RSv1 cohort at 150 records and the RSv2 cohort at 60 records; no formal statistical sample size calculation was performed. We quantified the precision of agreement estimates using bootstrap 95% confidence intervals.

### 2.2. Classification framework and reviewers

We used the four-layer, 19-category HFACS framework, with assessments performed by five physicians and one nurse who received common written instructions and calibration. The four layers were unsafe acts (L1), preconditions for unsafe acts (L2), unsafe supervision (L3), and organisational influences (L4); they were not treated as an ordinal severity scale [2,3]. We added three non-HFACS categories: patient condition (CoP), OTHER, and INAPPROPRIATE. Human classification used all 22 categories, whereas AI-to-reference comparisons in RSv1 combined the three non-HFACS categories, yielding 20 categories. Agreement was assessed at detailed-category, layer, and binary HFACS/non-HFACS levels.

The reviewers had a median of 8 years of clinical experience (range, 2–15 years), and none had used HFACS before the study. Before assessment, common written instructions and a calibration meeting addressed analysability and classification criteria. The same three reviewer pairs worked across all stages. For RSv1 and RSv2 construction, each reviewer assessed the assigned records independently before pair consensus. Category definitions and details of reviewer preparation are provided in the supplementary materials.

### 2.3. RSv1: Evaluation of HFACS classification of AI-generated candidates

We evaluated agreement in HFACS classification of AI-generated candidate factors both between reviewers and between AI predictions and the consensus reference standard, RSv1. Using a workflow developed on the separate 100-record sample, Kimi K2.5 generated FTA structures and HFACS classifications from the three narrative fields. FTA represented events and contributing factors in a tree linked by AND/OR gates; terminal candidates were termed leaf nodes. Contributing factors were actions, events or conditions judged from report narratives to contribute to the incident. Relationships were directed links representing how an event or factor was judged to contribute to another or to the incident. Both were inferred from the narratives rather than treated as established causation.

Reviewers independently assessed the narratives and AI candidates, making separate decisions on node retention and classification. After clarification of coding rules, they reassessed leaf nodes initially labelled L1 by either reviewer. The primary inter-reviewer analysis excluded each pair’s first 10 records, which coincided with calibration and procedural refinement, and included jointly retained leaf nodes from records both reviewers considered analysable. Detailed-category Cohen’s kappa was the primary measure; observed agreement, AC1, and layer-level and binary agreement were secondary measures.

Pair consensus determined retained candidates and their classifications to create RSv1. AI classification was evaluated on consensus-retained leaf nodes with one reference label, including non-HFACS nodes. In a later classification reanalysis, the 150 records were divided into development, model/prompt selection, and final evaluation sets of 50 each; the selection and final evaluation sets did not overlap. GPT-5.6 Sol and the selected classification prompt were frozen before final evaluation. Kimi K2.6 received the unchanged prompt for secondary evaluation. Detailed-category Cohen’s kappa was the primary measure; observed agreement, AC1, and layer-level and binary agreement were secondary measures. Target values were at least 0.6 for kappa and AC1. Candidate generation, reassessment, data splitting and sensitivity analyses are detailed in Supplementary Methods S2a, S4 and S5; model execution information is summarised in Supplementary Table SM2.

### 2.4. Evaluation of the added value of FTA

We assessed the added value of FTA by comparing structure-based category rankings with factor-frequency rankings and FTA-based classification with direct classification without FTA. These analyses included 87 records rated analysable in RSv1 after excluding each pair’s first 10 records. Tree regeneration and direct classification both used Kimi K2.5 with the event description and background as inputs; improvement measures were excluded.

First, we compared structure-based and factor-frequency category rankings from the same regenerated trees. Structure-based rankings counted records in which setting all leaves assigned to a category to false made the top event false under the generated AND/OR logic; frequency rankings counted records containing that category.

Reference categories comprised retained RSv1 HFACS leaf categories supported by verbatim improvement-measure quotations, supplemented by categories from reviewer-added factors and deduplicated within each report. Recall at five measured the proportion of reference report-category occurrences covered by the five highest-ranked categories. Corpus-wide scale-up required a positive lower confidence bound for the difference in recall at five, stable top-five selection, a top five different from factor-frequency rankings, and no detected improvement-input leakage.

We then conducted an exploratory comparison of FTA-based and direct multi-label classification. This comparison used report-level reference category sets derived from all retained RSv1 HFACS leaves and reviewer-added factors, without restriction to improvement quotations. We calculated record-averaged precision and recall and pooled micro-F1, with separate evaluation of combined L3/L4 performance. Ranking calculations, reranking within bootstrap samples and aggregation procedures are detailed in Supplementary Methods S6.

### 2.5. RSv2: Evaluation of contributing-factor extraction and classification

In a separate cohort of 60 records, reviewers independently extracted and classified contributing factors from report narratives without AI-generated candidates, and we evaluated agreement between their responses. This assessment was added after the initial RSv1 analyses, with factor-extraction agreement as the planned primary endpoint. Five separate pilot records were used to refine the task and interface and were excluded from formal performance estimates. The 60 records were selected using the reproducible procedure described in Section 2.1, without manual difficulty screening.

Each reviewer extracted factors without viewing AI candidates or their partner’s responses, supplied supporting quotations and classification labels where possible, and rated each record as analysable without uncertainty, analysable with uncertainty, or not analysable. Reviewers also recorded difficulty reasons, optional relationships between factors and evaluation time. Unknown actor or organisational scope was not, by itself, evidence for L3/L4.

Factors from the independent responses were matched one-to-one with AI assistance by their central proposition rather than identical wording, and extraction agreement was assessed using micro-F1. Matching was not independently verified by humans. The primary analysis included factors with an HFACS label or an indeterminate classification and excluded factors labelled only as non-HFACS. We also assessed classification agreement for matched factors and agreement on relationships between factors. Pairs subsequently adopted, merged or revised factors to create RSv2, and we summarised analysability, factor classifications and resolution status.

We additionally performed exploratory AI classification of all 110 RSv2 factors using the frozen GPT-5.6 Sol configuration. Inputs comprised factor texts and the original three narrative fields; reference labels, actor fields and resolution status were withheld. We calculated the proportion of predictions matching at least one reference label and detailed-category and layer-level kappa for factors with a single HFACS label. Non-HFACS predictions were retained as disagreements. Response versions, matching rules, multiple-label and missing-data handling, sensitivity analyses and resolution-status definitions are detailed in Supplementary Methods S7 and the related supplementary analyses.

### 2.6. Reassessment with additional public information

For the 60 RSv2 records, we evaluated changes in reviewers’ analysability judgements and HFACS classifications of existing factors after reassessment with additional public information. Reviewers independently revisited their saved responses alongside available public fields, including involved professions, event location and circumstances, and reporter-assigned contributing factors. Reporter-assigned factors were presented as reporter judgements, not HFACS ground truth.

Factor sets were fixed while reviewers confirmed or changed analysability and existing classification labels. They recorded information supporting changes and remaining uncertainties; exceptional factor additions or revisions were recorded as notes. No further consensus round was requested. Reassessment was sequential and non-blinded, with previous responses available.

The primary measures were within-reviewer analysability transitions and inter-reviewer linear weighted kappa for analysability. Classification changes and layer-level agreement were secondary measures. Before-and-after comparisons used the baseline responses actually presented during reassessment. Inter-reviewer classification comparisons included matched factors classified by both reviewers at both time points. Factor eligibility, multiple-label handling, layer-set agreement and evaluation-time summaries are detailed in Supplementary Methods S8.

### 2.7. Statistical analysis

We assessed classification and factor-extraction agreement primarily using kappa and micro-F1, respectively, and calculated 95% confidence intervals using record-cluster bootstrap resampling stratified by reviewer pair. We used 10,000 resamples and percentile intervals [15], preserving each pair’s record count and resampling all analysed nodes, factors and relationships within each record together. Paired FTA comparisons and fixed-factor before-and-after comparisons used the same resampled records for both methods or stages.

HFACS classifications were treated as nominal, and linear weighted kappa was used only for ordered analysability responses. Missing labels were retained and analysis-specific denominators were reported. We compared factor agreement by reported actor, role or scope uncertainty and assessed the association between record-level factor F1 and mean recorded reviewer time using Spearman’s correlation. Formulas, resampling settings and sensitivity analyses are detailed in Supplementary Methods S4–S8. Analyses used Python 3.12 and 3.14, with versioned code and software-environment records retained for reproducibility [16].

### 2.8. Ethical considerations

This study used only publicly available secondary data without personally identifiable information and collected no additional data from individuals. Institutional ethical review and informed consent were therefore not required. Data were accessed and used in accordance with the JCQHC terms governing use of the Project to Collect Medical Near-Miss/Adverse Event Information.

## 3. Results

The study included 150 records in the RSv1 cohort and 60 new records in the RSv2 cohort (Figure 1).

The RSv1 cohort yielded 300 reviewer responses and 2,073 original FTA nodes. RSv1 rated 112/150 records (74.7%) as analysable and 38/150 (25.3%) as not analysable. Among records 11–50 assigned to each pair, 81 records were considered analysable by both reviewers and entered the inter-reviewer classification analysis; 87 records were rated analysable in RSv1 and entered the FTA added-value analyses. In the later final AI classification evaluation, 37 of the 50 allocated records were analysable and contributed 306 leaf nodes. All 60 RSv2 records had two saved reviewer responses and were included in the additional-public-information reassessment.

### 3.1. Inter-reviewer classification agreement in RSv1

The primary inter-reviewer classification analysis included 81 records and 731 jointly retained leaf nodes and used post-L1-reassessment labels. Pooled kappa was 0.503 (95% CI, 0.456 to 0.548) for detailed 22-category classification, 0.529 (95% CI, 0.478 to 0.576) for HFACS layer classification, and 0.775 (95% CI, 0.716 to 0.825) for binary HFACS/non-HFACS classification.

The analysis included 24 records and 219 leaf nodes for AB, 29 records and 250 leaf nodes for CD, and 28 records and 262 leaf nodes for EF. Detailed-category kappa was 0.628 (95% CI, 0.551 to 0.695), 0.402 (95% CI, 0.307 to 0.484), and 0.488 (95% CI, 0.409 to 0.560), respectively. Observed agreement, AC1, and pair-specific estimates at each classification level are reported in Supplementary Table S1; sensitivity analyses are reported in Supplementary Table S3.

### 3.2. Added value of FTA

The FTA added-value analyses included 87 analysable RSv1 records. FTA-based structural rankings and factor-frequency rankings selected the same five categories; both achieved recall at five of 0.113 against improvement-related reference categories. The FTA-minus-frequency difference in recall at five was 0.000 (95% CI, −0.411 to 0.128). The criteria for a positive lower confidence bound and a top five different from frequency were not met; the stability criterion was met, and no improvement-input leakage flags were detected (Supplementary Table S4). Corpus-wide analysis did not proceed.

In the subsequent FTA-versus-direct-classification comparison using the same 87 records, the paired differences (FTA minus direct classification) were 0.086 for record-averaged precision (95% CI, 0.021 to 0.152) and −0.123 for record-averaged recall (95% CI, −0.195 to −0.050). The all-category micro-F1 difference was 0.032 (95% CI, −0.024 to 0.086). For L3/L4, micro-F1 was 0.325 for FTA and 0.457 for direct classification, a difference of −0.132 (95% CI, −0.237 to −0.028; Figure 2; Supplementary Table S5). Sensitivity analyses are reported in Supplementary Table S6.

**Figure 2.**
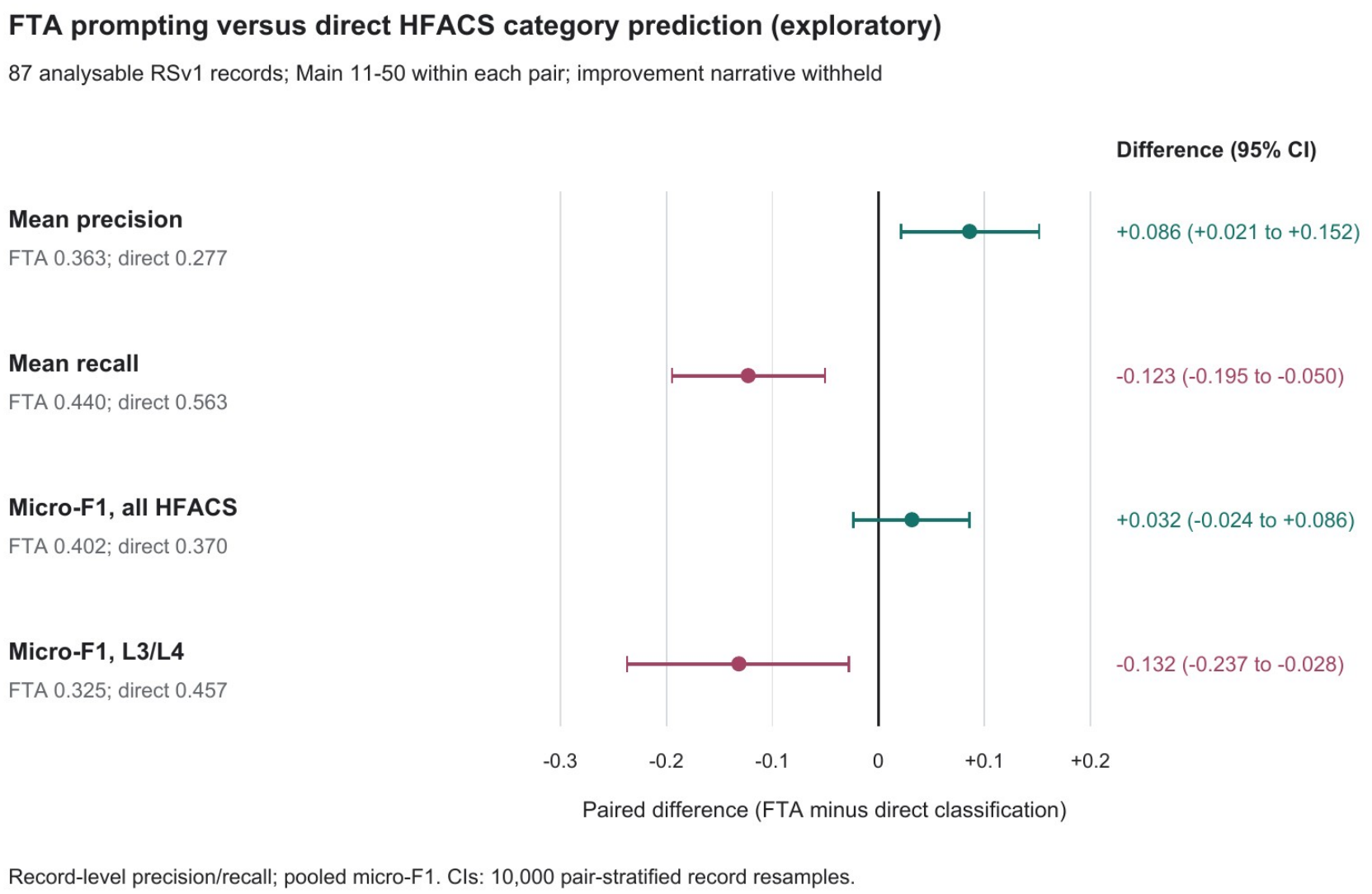
FTA versus direct HFACS category prediction under matched input conditions. Points show paired differences (fault tree analysis [FTA] minus direct classification) and 95% record-bootstrap confidence intervals in 87 analysable RSv1 cohort records from records 11–50 within each reviewer pair (labelled Main in the figure). Both methods used Kimi K2.5 with the event description and background as inputs; improvement measures were excluded. The reference targets comprised the RSv1-derived report-level set (Section 2.4). Precision and recall are record-level means; micro-F1 pools category counts. Intervals used 10,000 record-cluster bootstrap resamples stratified by reviewer pair. This exploratory comparison followed assessment of the criteria for corpus-wide analysis and does not measure clinical benefit. L3, unsafe supervision; L4, organisational influences.

### 3.3. AI classification agreement in RSv1

The final AI classification evaluation included 37/50 analysable records (74.0%), contributing 306 leaf nodes. Detailed-category kappa was 0.427 (95% CI, 0.359 to 0.493) for the selected GPT-5.6 Sol model and 0.499 (95% CI, 0.445 to 0.551) for the secondary Kimi K2.6 evaluation. Neither estimate reached the kappa target of 0.6.

Layer-level kappa was 0.437 for GPT-5.6 Sol and 0.514 for Kimi K2.6; binary kappa was 0.668 and 0.734, respectively (Table 1; Figure 3A). Detailed-category AC1 was 0.653 (95% CI, 0.576 to 0.720) and 0.676 (95% CI, 0.610 to 0.737), respectively; both point estimates exceeded the AC1 target of 0.6. Observed agreement and AC1 at all three classification levels, with their 95% confidence intervals, are reported in Supplementary Table S2.

**Table 1.** AI classification agreement on 306 RSv1 leaf nodes.

| **Model and evaluation role** | **Detailed kappa (95% CI)** | **Layer kappa (95% CI)** | **Binary kappa (95% CI)** |
|---|---|---|---|
| GPT-5.6 Sol, selected primary model | 0.427 (0.359 to 0.493) | 0.437 (0.365 to 0.509) | 0.668 (0.574 to 0.754) |
| Kimi K2.6, secondary evaluation | 0.499 (0.445 to 0.551) | 0.514 (0.453 to 0.573) | 0.734 (0.654 to 0.809) |

Both models were assessed on the same 37 analysable records and 306 leaves from the final 50-record set. Detailed classification included 19 HFACS categories and one combined non-HFACS category; layer classification included L1–L4 and non-HFACS. The Kimi evaluation was added after the selected model's final run. It did not replace the primary result.

**Figure 3.**
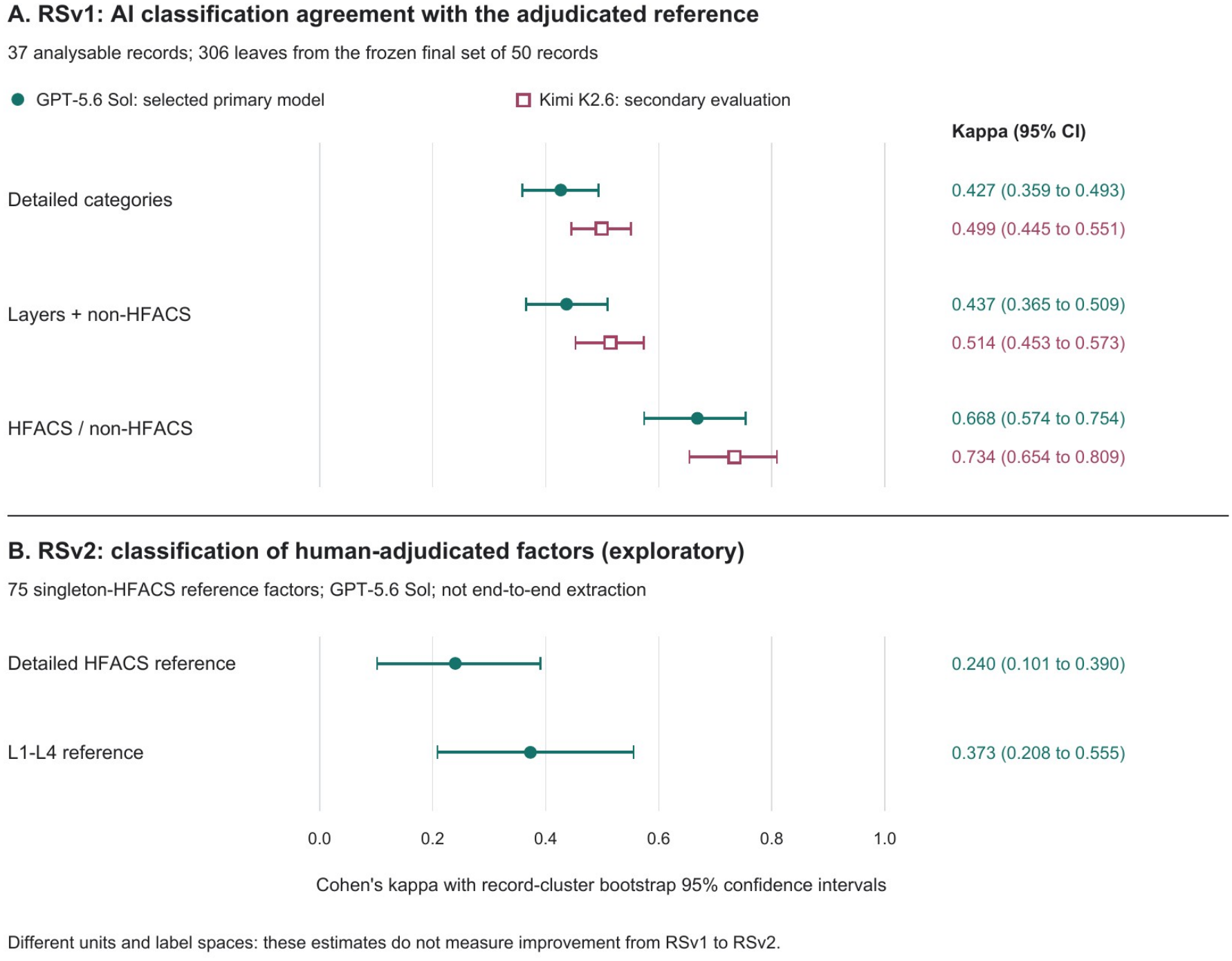
AI classification agreement with RSv1 and RSv2. Points show Cohen’s kappa and record-cluster bootstrap 95% confidence intervals stratified by reviewer pair. A: 306 leaves from 37 analysable RSv1 records (10,000 resamples); detailed and layer scoring included non-HFACS categories. GPT-5.6 Sol was primary and Kimi K2.6 secondary. B: AI classification of 75 fixed RSv2 factors with a single HFACS label (10,000 resamples); non-HFACS predictions were retained. Panel B is exploratory conditional classification, not end-to-end extraction. Cohorts, units and label spaces differ between panels. AI, artificial intelligence; HFACS, Human Factors Analysis and Classification System; RS, reference standard.

### 3.4. Agreement in contributing-factor extraction and the RSv2 dataset

All 60 records in the RSv2 cohort had two saved reviewer responses, yielding 248 extracted factors; 176 were eligible under the HFACS-or-indeterminate rule.

RSv2 contained 110 factors across 60 records. Record-level analysability was without uncertainty in 15/60 records (25.0%), with uncertainty in 23/60 (38.3%), and not analysable in 22/60 (36.7%). Fifteen factors had multiple classification labels, including 10 spanning multiple HFACS layers; five had no classification label. Human-assigned factor-level resolution status was source-confirmed for 48 factors, rule-adjudicated for one, unresolved for two, and missing for 59/110 (53.6%; Supplementary Table S8).

AI-assisted matching identified 37 eligible factor pairs, yielding micro-F1 of 0.420 (95% CI, 0.303 to 0.527; Table 2). Both reviewers extracted no eligible factors in 14/60 records (23.3%).

**Table 2.** Human factor-extraction agreement across 60 records in the RSv2 cohort.

| **Pair** | **Eligible factors reviewer 1 / 2** | **Matched factors** | **Micro-F1 (95% CI)** |
|---|---|---|---|
| AB | 23 / 29 | 9 | 0.346 (0.169 to 0.514) |
| CD | 36 / 33 | 14 | 0.406 (0.222 to 0.575) |
| EF | 22 / 33 | 14 | 0.509 (0.258 to 0.698) |
| Overall | 81 / 95 | 37 | 0.420 (0.303 to 0.527) |

Each pair assessed 20 different records. Factors with HFACS or indeterminate labels were eligible. Semantic correspondence was AI-assisted and had not been confirmed by humans. Bootstrap intervals captured record sampling variability conditional on those correspondences, not their potential errors.

In sensitivity analyses, micro-F1 was 0.315 using all factors and 0.416 when restricted to HFACS-labelled factors. Under the main eligibility rule, micro-F1 was 0.148 with strict quotation matching and 0.511 with partial overlap (Supplementary Table S7).

Among matched factor pairs, baseline exact-set agreement on recorded classification labels, including indeterminate, was 20/37 (54.1%). All 37 matched pairs had one label per reviewer, giving pooled nominal kappa of 0.446. Both reviewers assigned an HFACS layer in 32 pairs; exact layer-set agreement was 25/32 (78.1%), with pooled nominal kappa of 0.639. Confidence intervals are reported in Supplementary Results.

Exploratory inter-reviewer micro-F1 for relationships between contributing factors was 0.291: 23 links matched among 74 and 84 eligible links recorded by the two reviewers, respectively (Supplementary Results).

All saved quotations could be located in the supplied narratives for the 248 reviewer-extracted factors and 110 RSv2 factors. Quotations came exclusively from the improvement-measure field for 70/248 reviewer-extracted factors (28.2%) and 35/110 RSv2 factors (31.8%). Median saved reviewer evaluation time was 4.84 minutes (interquartile range [IQR], 2.24 to 11.80).

In the baseline exploratory analysis, factor micro-F1 was 0.453 in 34 records where either reviewer reported unclear actor, role, or scope, and 0.371 in the other 26. Across 46 records with defined factor F1 and recorded time, Spearman’s correlation between record-level factor F1 and mean recorded reviewer time was −0.001.

### 3.5. Reassessment with additional public information

All 120 reassessments were complete. Responses without uncertainty accounted for 45/120 (37.5%) before and 56/120 (46.7%) after reassessment; not-analysable responses accounted for 41/120 (34.2%) and 42/120 (35.0%), respectively (Figure 4). Nineteen responses moved to a more analysable state, seven to a less analysable state, and 94 retained their original rating.

**Figure 4.**
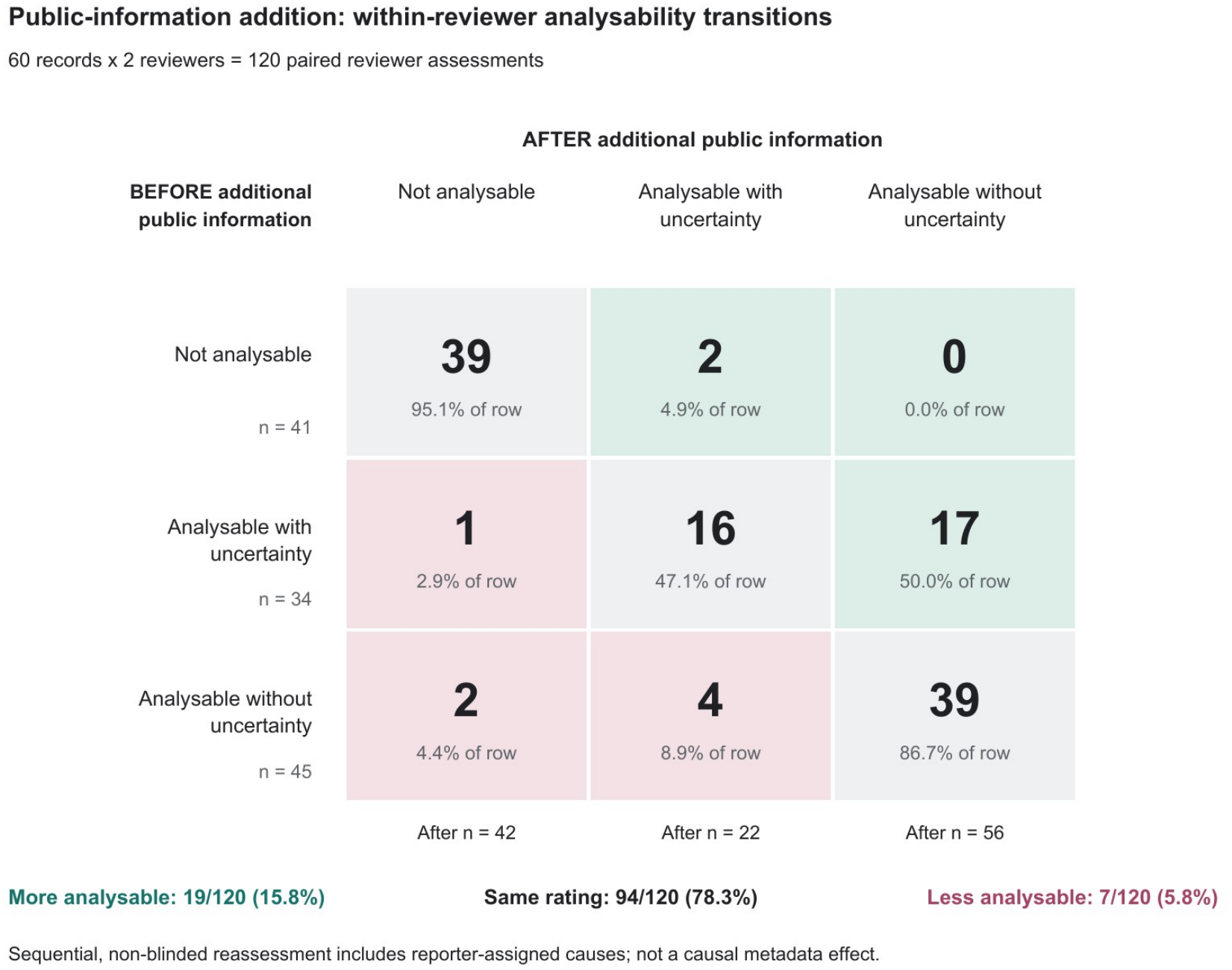
Analysability before and after additional public information. Cells show counts and row percentages of paired reviewer assessments. The matrix compares each reviewer’s rating before and after reassessment, comprising 120 paired assessments of 60 records by two reviewers per record. Green cells denote movement to a more analysable rating, grey cells an unchanged rating and rose cells a less analysable rating. Additional information included factual public fields and reporter-assigned cause judgements. Reassessment was sequential and non-blinded and could not isolate effects of metadata, rereading, prior exposure to RSv2 or reporter judgements. Rating changes do not establish correctness or improved inter-rater agreement.

Exact inter-reviewer analysability agreement was 31/60 records (51.7%) before and 26/60 (43.3%) after reassessment. Pooled linear weighted kappa was 0.386 (95% CI, 0.189 to 0.564) before reassessment and 0.203 (95% CI, −0.011 to 0.395) after reassessment. By pair, weighted kappa changed from 0.398 to 0.113 for AB, 0.348 to 0.245 for CD, and 0.400 to 0.388 for EF (Supplementary Table S9).

The response versions presented for reassessment contained 244 factors; four patient-condition-only factors in the independent-extraction versions were not present in this baseline (Supplementary Methods S7). Across 244 paired before-and-after assessments of existing factors by the same reviewer, recorded classification-label sets changed in 26 (10.7%). Six factors changed from a recorded classification to no classification (Supplementary Results, classification-change breakdown).

Among 32 matched factor pairs with HFACS layers assigned by both reviewers at both time points, exact inter-reviewer layer-set agreement changed from 25/32 (78.1%) to 21/32 (65.6%), whereas the proportion sharing at least one HFACS layer changed from 25/32 (78.1%) to 27/32 (84.4%). For the 26 matched factor pairs assigned a single layer by both reviewers at both time points, pooled nominal kappa remained 0.668 (Supplementary Table S10). Median reassessment activity time was 1.13 minutes (IQR, 0.46 to 2.97).

### 3.6. AI classification agreement in RSv2

AI classified all 110 RSv2 factor texts. Predictions matched at least one RSv2 label for 46/105 factors with any RSv2 label (43.8%) and 32/90 factors with at least one HFACS label in RSv2 (35.6%). Among 75 factors with a single HFACS label in RSv2, detailed-category agreement was 25/75 (33.3%), with kappa 0.240 (95% CI, 0.101 to 0.390); layer agreement was 40/75 (53.3%), with kappa 0.373 (95% CI, 0.208 to 0.555; Figure 3B).

## 4. Discussion

### 4.1. Principal findings

Our staged evaluations of public medical incident reports identified disagreement in factor extraction between reviewers and in HFACS classification both between reviewers and between AI predictions and the reference standards. Under the tested conditions, FTA did not establish added value over direct classification or factor-frequency ranking. In RSv1, neither AI model reached the detailed-category kappa target, and exploratory AI classification of RSv2 factors with a single HFACS label yielded a detailed-category kappa of 0.240. Reviewer factor extraction without AI-generated candidates also showed disagreement, and most RSv2 records were analysable with uncertainty or not analysable. Before and after reassessment with additional public information, responses without uncertainty accounted for 37.5% and 46.7%, exact inter-reviewer analysability agreement was 51.7% and 43.3%, and pooled linear weighted kappa was 0.386 and 0.203, respectively.

### 4.2. Interpretation and comparison with previous studies

Disagreement in factor extraction between reviewers indicates that the challenge extends beyond AI classification performance. Ebara et al. reported high inter-reviewer agreement in sentence-level P-mSHELL classification of public medication-related reports and good BERT classification performance for some categories [7]. They excluded 20 of 1,212 initially retrieved reports (1.7%) because they lacked background-factor descriptions. After segmenting the background-factor field, they also excluded sentences with missing or unclear background information or descriptions of post-incident actions, retaining 5,058 sentences for classification [7]. In our RSv2 cohort, reviewers extracted specific factors without AI-generated candidates from reports selected without manual difficulty screening, and disagreement was observed at this stage. Smits et al. evaluated root-cause identification and classification separately, while Wang et al. evaluated model classification of expert-extracted factor statements, also distinguishing factor identification from classification [6,13]. Thus, classification performance on prepared inputs does not establish whether the same contributing factors can be identified from collected reports. Our findings indicate the need to consider consistency in factor identification and the reporting information that supports it, alongside improvements in classification technology. This supports reporting-system designers and operators strengthening documentation support so that the circumstances and context needed to interpret contributing factors and relationships can be recorded.

Limited contextual detail may partly explain the assessment difficulties. Nuckols et al. identified insufficient narrative detail for specific classification of provider factors, and Cohen et al. highlighted limited contextual information for identifying supervisory and organisational factors [4,17]. Our assessment difficulties are consistent with these observations, and Ebara et al.’s exclusion of sentences with unclear background information further highlights the importance of considering input conditions when interpreting classification performance [7]. Although ambiguous descriptions, extraction criteria and reviewer judgement remain possible explanations, processes for recording the circumstances and context needed to interpret contributing factors and relationships also warrant attention alongside analytical methods. Cohen et al.’s Systems Approach For Event Reporting (SAFER) programme demonstrated the feasibility and acceptability of training nurses to organise event narratives and background information using Situation, Background, Assessment, Recommendation (SBAR) and describe contributing factors using the Systems Engineering Initiative for Patient Safety (SEIPS) model [18]. Such documentation support offers a practical example for reporting systems, but its contribution to agreement in factor extraction and classification requires evaluation.

### 4.3. Implications for reporting systems

Organisations that operate incident reporting systems or develop reporting guidance could strengthen reporting forms, guidance and reporter education to help reporters document the circumstances needed to interpret contributing factors and relationships. Rather than simply requesting more detail, such support could use examples and prompts to help reporters describe the sequence of events and the surrounding work-system conditions [18]. In the US railway sector, the Federal Railroad Administration (FRA) funds the Confidential Close Call Reporting System (C3RS), with NASA receiving and analysing reports as an independent third party. The system uses reporting examples and guidance within reporting forms. When reports lack detail, NASA analysts contact reporters by telephone to obtain additional information, supported by standardised questions tailored to the event type [19]. Healthcare reporting systems could support documentation of the circumstances and context needed to interpret contributing factors and relationships through reporting examples, guidance within reporting forms and, where needed, follow-up with reporters. The purpose is to provide a clearer basis for interpreting contributing factors and relationships, not to require reporters to establish causation. The usefulness of this support should be evaluated in terms of its contribution to factor extraction and classification, rather than increases in narrative length.

These implications may also apply to reporting systems in other countries that combine structured fields with narratives. The combination of structured items and narrative descriptions in JCQHC reports is not unique to Japan. It is also used in the US Agency for Healthcare Research and Quality (AHRQ) Common Formats and Finland’s HaiPro system [12,20,21]. The World Health Organization (WHO) Minimal Information Model for Patient Safety Incident Reporting and Learning Systems similarly combines standardised structured information with free-text incident descriptions to support comparison across countries [22]. In HaiPro, Holmström et al. found disagreement between local classifiers and a researcher assessing the same medication-error reports, with the researcher identifying more contributing factors [21]. Although they compared routine classifications with researcher reassessment, whereas we assessed agreement between paired reviewers using HFACS on public reports, both findings suggest that structured categories alone do not eliminate differences in narrative interpretation. Assessing agreement in identifying contributing factors from narratives is therefore relevant to similar reporting systems in other countries.

### 4.4. Limitations

There are several limitations. First, we did not compare public records with complete original hospital reports. Context omitted during public release could therefore be mistaken for information never recorded, overstating the contribution of the original information-collection process to assessment difficulties. Second, despite common written instructions, practice assessments and calibration, reviewers had no prior HFACS experience. If unfamiliarity contributed to disagreement, attributing that disagreement to report content could overstate the contribution of insufficient information. Third, access to AI-generated candidates and classifications during RSv1 construction may have increased AI-to-reference agreement through anchoring. AI-assisted factor matching in the RSv2 cohort was not independently verified by humans; missed matches could lower micro-F1 and overstate disagreement in factor extraction, whereas incorrect matches could raise micro-F1 and understate disagreement. The effects of anchoring and matching errors were not quantified; sensitivity analyses examined dependence on eligibility and matching criteria, not matching accuracy. Finally, rereading and exposure to RSv2 during reassessment could influence responses and inter-reviewer agreement independently of the additional public information, confounding interpretation of its effects. Factor sets were fixed, so potential effects on identifying new factors were not assessed. Overall, these limitations prevent identification of the original information-collection process as the main cause of the observed assessment difficulties.

### 4.5. Conclusions

In evaluations of public medical incident reports, reviewers disagreed on contributing-factor extraction, and neither AI model met the detailed-category kappa target for Human Factors Analysis and Classification System (HFACS) classification against the first reviewer-consensus reference standard (RSv1). Difficulty discerning relevant circumstances and context from the available reports may have contributed to these assessment difficulties. Organisations that operate incident reporting systems or develop reporting guidance could strengthen reporting forms, guidance and reporter education to help reporters document the circumstances and context needed to interpret contributing factors and relationships.

## Data Availability

The companion repository contains pseudonymised numeric and categorical analysis inputs, saved aggregate results, versioned analysis scripts, prompt templates, and reproduction instructions [16]. Source incident narratives and information identifying individual reviewers are not included.

https://github.com/FuyutoMiyake/hfacs-incident-report-reproducibility/tree/a1e27b59adcd02741c4efd026f48bf2c0be42d9a

https://github.com/FuyutoMiyake/hfacs-incident-report-reproducibility/blob/f756c8a17d77525f5625e1f5d3c47b7cc2e775a7/supplements/README.md

## Use of artificial intelligence

ChatGPT and Codex (OpenAI), Claude (Anthropic), and Google Deep Review were used to assist with manuscript preparation. ChatGPT and Codex were also used to assist with literature searches and analysis code development. All AI-generated suggestions were reviewed and verified by the authors, who take responsibility for the accuracy and integrity of the content.

## References

1. M.F. Peerally, S. Carr, J. Waring, G. Martin, M. Dixon-Woods, A content analysis of contributory factors reported in serious incident investigation reports in hospital care, Clin. Med. (Lond.) 22 (2022) 423–433. 10.7861/clinmed.2022-0042.

2. D.A. Wiegmann, S.A. Shappell, A Human Error Approach to Aviation Accident Analysis: The Human Factors Analysis and Classification System, Routledge, 2017. 10.4324/9781315263878.

3. T. Diller, G. Helmrich, S. Dunning, S. Cox, A. Buchanan, S. Shappell, The Human Factors Analysis Classification System (HFACS) applied to health care, Am. J. Med. Qual. 29 (2014) 181–190. 10.1177/1062860613491623.

4. T.N. Cohen, S.E. Francis, D.A. Wiegmann, S.A. Shappell, B.L. Gewertz, Using HFACS-Healthcare to identify systemic vulnerabilities during surgery, Am. J. Med. Qual. 33 (2018) 614–622. 10.1177/1062860618764316.

5. A.J. Jinia, K. Chapman, S. Liu, C. Della Biancia, E. Hipp, E. Lin, R. Moulder, D. Parikh, J. Cordero, C. Pistone, M. Gil, J. Ford, A. Li, J.M. Moran, Artificial intelligence-based incident analysis and learning system to enhance patient safety and improve treatment quality, npj Digit. Med. 9 (2026) 254. 10.1038/s41746-026-02390-2.

6. Y. Wang, L. Bowditch, C. Molloy, Y. Yu, P. Hibbert, F. Magrabi, Fine-tuning and evaluating large language models for patient safety tasks: classification of contributing factors in incident reports, J. Am. Med. Inform. Assoc. 33 (2026) 1494–1503. 10.1093/jamia/ocag068.

7. S. Ebara, H. Kizaki, Y. Takamatsu, S. Watabe, S. Imai, S. Yada, E. Aramaki, O. Yasumuro, R. Funakoshi, S. Hori, Automatic extraction of multiple contributing factors in medication-related incidents based on the P-mSHELL model: development and evaluation of a BERT-based multilabel classifier, BMJ Health Care Inform. 33 (2026) e102260. 10.1136/bmjhci-2026-102260.

8. J. Kottner, L. Audigé, S. Brorson, A. Donner, B.J. Gajewski, A. Hróbjartsson, C. Roberts, M. Shoukri, D.L. Streiner, Guidelines for Reporting Reliability and Agreement Studies (GRRAS) were proposedJ. Clin. Epidemiol. 64 (2011) 96–106. 10.1016/j.jclinepi.2010.03.002.

9. Japan Council for Quality Health Care, Project to Collect Medical Near-Miss/Adverse Event Information: Outline. https://www.med-safe.jp/contents/english/index.html (accessed 11 October 2026).

10. Japan Council for Quality Health Care, Frequently asked questions: participation and reporting. https://www.med-safe.jp/contents/faq/index.html (accessed 14 October 2026).

11. Japan Council for Quality Health Care, QI-01 Registered institutions as of 31 March 2025. https://www.med-safe.jp/contents/report/html/shihannki/2025/1Q/TTL372_QI-01.html (accessed 14 October 2026).

12. Japan Council for Quality Health Care, Public case search: information on published cases. https://www.med-safe.jp/mpsearch/SearchReport.action (accessed 11 October 2026).

13. M. Smits, J. Janssen, R. de Vet, L. Zwaan, D. Timmermans, P. Groenewegen, C. Wagner, Analysis of unintended events in hospitals: inter-rater reliability of constructing causal trees and classifying root causes, Int. J. Qual. Health Care 21 (2009) 292–300. 10.1093/intqhc/mzp023.

14. J.K. O’Hara, C. Reynolds, S. Moore, G. Armitage, L. Sheard, C. Marsh, I. Watt, J. Wright, R. Lawton, What can patients tell us about the quality and safety of hospital care? Findings from a UK multicentre survey study, BMJ Qual. Saf. 27 (2018) 673–682. 10.1136/bmjqs-2017-006974.

15. T.C. Hesterberg, What teachers should know about the bootstrap: resampling in the undergraduate statistics curriculum, Am. Stat. 69 (2015) 371–386. 10.1080/00031305.2015.1089789.

16. FuyutoMiyake, HFACS incident-report analysis: computational companion [software and data], GitHub, version a1e27b59 (2026). https://github.com/FuyutoMiyake/hfacs-incident-report-reproducibility/tree/a1e27b59adcd02741c4efd026f48bf2c0be42d9a (accessed 9 November 2026).

17. T.K. Nuckols, D.S. Bell, S.M. Paddock, L.H. Hilborne, Contributing factors identified by hospital incident report narratives, Qual. Saf. Health Care 17 (2008) 368–372. 10.1136/qshc.2007.023721.

18. T.N. Cohen, T.K. Nuckols, C.T. Berdahl, E.G. Seferian, S.G. McCleskey, A.J. Henreid, D.W. Leang, M.A. Lupera, B.L. Coleman, Training hospital nurses to write detailed narratives and describe contributing factors in incident reports: the SAFER Education Program, Jt. Comm. J. Qual. Patient Saf. 51 (2025) 305–311. 10.1016/j.jcjq.2025.01.002.

19. J. Multer, H. Safar, H. Howarth, J. Gissel, T. Parker, K. Stowe, Evaluating Processes, Outcomes, and Sustainability for FRA’s Confidential Close Call Reporting System (C3RS), Federal Railroad Administration, Washington, DC, 2024. Report DOT/FRA/ORD-24/45. https://rosap.ntl.bts.gov/view/dot/78972.

20. Agency for Healthcare Research and Quality, Common Formats for Event Reporting - Hospital Version 2.0aR: Event Description, Generic - National Collection (Core), March 2025. https://www.psoppc.org/api/dlms/downloadDocument?groupId=2589.

21. A.-R. Holmström, R. Järvinen, R. Laaksonen, T. Keistinen, P. Doupi, M. Airaksinen, Inter-rater reliability of medication error classification in a voluntary patient safety incident reporting system HaiPro in Finland, Res. Social Adm. Pharm. 15 (2019) 864–872. 10.1016/j.sapharm.2018.11.013.

22. World Health Organization, Minimal Information Model for Patient Safety Incident Reporting and Learning Systems: User Guide, WHO, Geneva, 2016, WHO/HIS/SDS/2016.22. https://www.who.int/publications/i/item/WHO-HIS-SDS-2016.22.

